# Time to Medication in School-Age Children with ADHD: Assessing the Effect of Sociodemographic and Clinical Factors

**DOI:** 10.1101/2025.02.21.25322686

**Authors:** Sushma Kasinathan, Lynne C. Huffman, Ingrid Luo, Yair Bannett

**Affiliations:** Division of Developmental-Behavioral Pediatrics, Department of Pediatrics, Stanford University School of Medicine, Palo Alto, California; Quantitative Sciences Unit, Stanford University School of Medicine, Palo Alto, California

**Author notes:** Address correspondence to: Yair Bannett, Developmental-Behavioral Pediatrics, Stanford University School of Medicine, 3145 Porter Drive, Palo Alto, CA, 94304.

**Keywords:** Attention-Deficit/Hyperactivity Disorder, Developmental-Behavioral Pediatrics, Sociodemographic Factors, Clinical Factors, Healthcare Disparities

## Abstract

**Objective:** To assess the timing of attention-deficit/hyperactivity disorder (ADHD) medication prescriptions in relation to the ADHD diagnostic visit in school-aged children seen in developmental-behavioral pediatrics (DBP) clinics and to investigate which sociodemographic and clinical factors affect the timing of medication prescription.

**Method:** We retrospectively analyzed electronic health records from Stanford’s DBP clinics of children 6-12-years-old seen between 2016 and 2024. ADHD cohort included patients with an ICD-10 ADHD diagnosis and with ≥1 month follow-up. Andersen Health Care Utilization model informed selection of predisposing (age, sex, race/ethnicity), enabling (insurance, clinician type), and need (ADHD subtype, comorbidities) factors. Primary outcome: Time from first ADHD diagnosis to first ADHD medication prescription within 1 year of diagnosis. Cox regression model assessed associations between sociodemographic and clinical factors and study outcome.

**Results:** Of 823 patients with ADHD, 623 (75.7%) were male; average age at first ADHD diagnosis was 8.2 years (SD=1.5). Of 823 patients, 484 (58.8%) were prescribed medications within 1 year of diagnosis. The average number of days from ADHD diagnosis to first prescription was 51 days (median=4). Longer time to medication prescription was associated with Asian race/ethnicity (predisposing), psychologist clinician (enabling), 1-2 or ≥3 comorbidities (need). Shorter time to prescription was associated with older patient age (predisposing), ADHD combined and hyperactive/impulsive subtypes (need).

**Conclusion:** Asian race/ethnicity and, unexpectedly, multiple comorbidities, were associated with later ADHD medication prescription by DBPs in school-age children. Further investigation is necessary to understand patient, family, and clinician factors that influence medication initiation in these patient subgroups.

## INTRODUCTION

Attention-deficit/hyperactivity disorder (ADHD) is the most common pediatric neurodevelopmental disorder affecting children in the United States.^1^ Population-based studies have shown that ADHD often persists into adulthood and is associated with life-long adverse outcomes including poor mental health, academic and employment problems, substance abuse, incarceration, and mortality.^2–5^ Medication treatment improves core symptoms of ADHD and may also carry long-term benefits, such as reduced risk of associated mental health problems.^6–8^

Clinical practice guidelines by the American Academy of Pediatrics (AAP) and the Society for Developmental and Behavioral Pediatrics (SDBP) recommend FDA-approved medications for ADHD as the first-line treatment in school-age children (ages 6 to 12 years), in combination with parent training in behavior management and/or school-based interventions.^9,10^ However, many children with ADHD do not receive recommended treatments.^11^ The 2016 United States National Survey of Children’s Health revealed that approximately one-fourth of children aged 2-17 years with ADHD had never received any type of evidence-based treatment.^12^

Studies have identified patient clinical presentation and sociodemographic characteristics as important factors that affect parent report of diagnosis and treatment of children with ADHD.^13^ Other studies have described the contribution of health services factors, such as access to prescribing providers, insurance type, and availability of follow-up appointments to receipt of ADHD-related health care but not particularly in DBP subspecialty settings.^14,15^ Furthermore, prior studies have not applied a comprehensive conceptual model to investigate ADHD-related health care.

In this study, our objectives were (1) to assess the timing of ADHD medication initiation in school-age children seen in DBP clinics and (2) to identify specific patient-level and clinician-level facilitators and inhibitors associated with the timing of ADHD medication treatment. We used an established conceptual framework, the Andersen’s Behavioral Model of Health Service.^16^ This framework allowed us to assess the influence of pre-established categories, including predisposing, enabling, and need factors. Predisposing factors are factors which patients came to the study with, such as social and demographic characteristics, enabling factors are those that influence patients’ access to care, and need factors are those that would motivate patients to get care as determined by themselves and their providers. We hypothesized that non-White race/ethnicity (predisposing) and no comorbidities (need) would be associated with longer time to prescription.

## METHODS

### Study Population

In this retrospective cohort study, we extracted electronic health record (EHR) data from clinical encounters of children aged 6-12 years seen at Stanford’s Developmental-Behavioral Pediatrics (DBP) clinics between January 1, 2016, and June 30, 2024. We defined our study cohort of children with ADHD based on ICD-10 visit diagnosis codes for ADHD, including F90.0, F90.1, F90.2, F90.8, and F90.9 (excluding symptom-level descriptors, such as hyperactivity). We included patients with at least two visits at a DBP clinic and a minimum of one month between their first ADHD diagnostic visit and their last DBP visit. Exclusion criteria included: (1) presence of an autism spectrum disorder diagnosis prior to or concurrent with the first ADHD diagnostic visit, (2) ADHD diagnosis at DBP clinic before age 6, and (3) prescription of ADHD medications (stimulants and non-stimulants) by DBP prior to the diagnostic visit. ADHD medication prescription was identified by Observational Medical Outcomes Partnership (OMOP) codes for methylphenidate (705944), dexmethylphenidate (731533), amphetamine (714785), dextroamphetamine (719311), lisdexamfetamine (709567), guanfacine (1344965), clonidine (1398937), atomoxetine (742185), or any descendant concept. The final cohort consisted of 823 patients (**Figure 1** illustrates the study flowchart).

**Figure 1.**
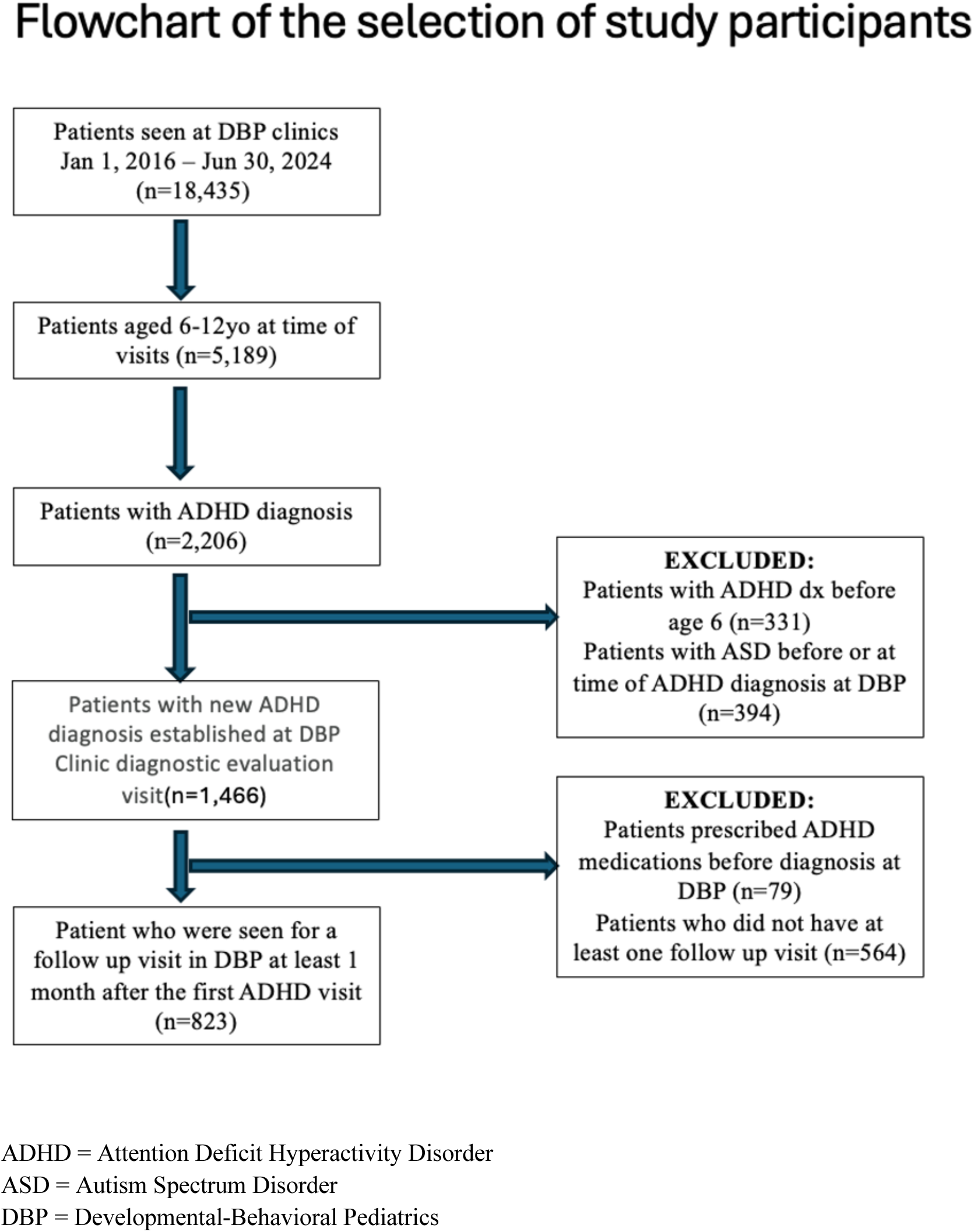
Study Flowchart

### Data Source

Electronic health records were extracted from STAnford medicine Research data Repository (STARR), which uses the OMOP common data model. This database provided de-identified patient records, including structured data of patient sociodemographic information and clinical data from visits (e.g., diagnosis, prescription). We received approval from Stanford’s Institutional Review Board to conduct this study.

### Outcome Measure

The primary outcome was time from initial ADHD diagnostic visit to first prescription of any ADHD medication. Patients were censored one year after their diagnostic visit if no ADHD medication was prescribed during this period.

### Predictors

Andersen’s Behavioral Model of Health Services Use provides a theoretical structure for identifying the factors that affect an individual’s use of health services.^16^ We used this model to identify various predisposing, enabling, and need factors that could be associated with our study outcome - time from ADHD diagnosis to medication prescription.

In alignment with this model, we selected all available data in the EHR that fit any of the conceptual model categories. Predisposing sociodemographic factors included patient age (at the diagnostic visit), sex (male or female), and race/ethnicity (Asian, Black or African American, Hispanic, Other, Unknown, or White). Enabling factors included provider type (physician, nurse practitioner, or psychologist) and insurance type (private, public, or missing). Need factors included ADHD subtype (combined, predominantly hyperactive, predominantly inattentive, or unspecified), and associated developmental-behavioral comorbid conditions (e.g., language delay/disorder, anxiety), classified as none, 1-2, or ≥3 comorbidities (see **eSupplement** for list of conditions and codes).

### Statistical Analyses

Descriptive statistics (n, proportions, means ± SD, medians, interquartile ranges) were used to summarize patient characteristics, stratified by whether patients received an ADHD medication prescription within one year of diagnosis or not. Group comparisons were performed using Kruskal-Wallis tests for continuous variables and Fisher’s exact tests for categorical variables. Time-to-event analyses applied stratified cumulative incidence functions by race/ethnicity and Cox proportional hazards models to assess predictors of time to first ADHD medication prescription. Models were adjusted for patient observation period (i.e., number of days from first to last visit at DBP clinic). The proportional hazards assumption was tested using Schoenfeld residuals. Statistical significance was assessed using a two-tailed p-value < 0.05. All statistical analyses were conducted using R version 4.3.2.

## RESULTS

**Table 1** displays the characteristics of the study cohort. Of the 823 patients with an initial ADHD diagnosis given at DBP clinics between ages 6-12 years, 369 (76.2%) were male, 291 (35.4%) were White, and 236 (28.7%) were Hispanic. The average age at first ADHD diagnosis was 8.2 years (SD 1.52). Of 823 patients with ADHD, 484 (58.8%) were prescribed an ADHD medication within 1 year of diagnosis. As shown in **Table 1**, compared to patients who were not prescribed a medication within a year of diagnosis, patients prescribed medications were older (mean age 8.3 vs. 8.0 years, p=0.008), less frequently diagnosed by a psychologist (6.0% of patients vs. 14.2%, p=0.004), more frequently had no comorbid conditions (26.4% of patients vs. 17.1%, p=0.029), and more frequently had an ADHD combined subtype diagnosis (55.4% of patients vs. 45.4%, p=0.005). Patient sex, race/ethnicity, and insurance type did not differ between those prescribed and not prescribed medications.

**Table 1.** ADHD Patient demographics.

|  | **Prescribed ADHD medication**<br>**within 1 year** |  | **Overall** | **P value** |
| --- | --- | --- | --- | --- |
|  | Yes (N=484) | No (N=339) | N = 823 |  |
| **PREDISPOSING FACTORS** |  |  |  |  |
| Sex |  |  |  | 0.918 |
| Male | 369 (76.2%) | 254 (74.9%) | 623 (75.7%) |  |
| Female | 115 (23.8%) | 85 (25.1%) | 200 (24.3%) |  |
| Age at diagnosis |  |  |  | 0.008 |
| Median (Q1, Q3) | 8.10 (7.16,<br>9.45) | 7.60 (6.79,<br>9.07) | 7.91 (6.94,<br>9.31) |  |
| Race/Ethnicity |  |  |  | 0.311 |
| Asian | 36 (7.4%) | 42 (12.4%) | 78 (9.5%) |  |
| Black or African American | 8 (1.7%) | 8 (2.4%) | 16 (1.9%) |  |
| Hispanic | 137 (28.3%) | 99 (29.2%) | 236 (28.7%) |  |
| Other | 3 (0.6%) | 5 (1.5%) | 8 (1.0%) |  |
| Unknown | 112 (23.1%) | 82 (24.2%) | 194 (23.6%) |  |
| White | 188 (38.8%) | 103 (30.4%) | 291 (35.4%) |  |
| **ENABLING FACTORS** |  |  |  |  |
| Insurance plan |  |  |  | 0.746 |
| Private | 335 (69.2%) | 227 (67.0%) | 562 (68.3%) |  |
| Public | 139 (28.7%) | 106 (31.3%) | 245 (29.8%) |  |
| Missing | 10 (2.1%) | 6 (1.8%) | 16 (1.9%) |  |
| Provider at diagnosis |  |  |  | 0.004 |
| Physician | 441 (91.1%) | 280 (82.6%) | 721 (87.6%) |  |
| Nurse Practitioner | 14 (2.9%) | 11 (3.2%) | 25 (3.0%) |  |
| Psychologist | 29 (6.0%) | 48 (14.2%) | 77 (9.4%) |  |
| **NEED FACTORS** |  |  |  |  |
| Comorbid condition |  |  |  | 0.029 |
| None | 128 (26.4%) | 58 (17.1%) | 186 (22.6%) |  |
| 1-2 | 302 (62.4%) | 231 (68.1%) | 533 (64.8%) |  |
| ≥3 | 54 (11.2%) | 50 (14.7%) | 104 (12.6%) |  |
| ADHD Diagnosis Subtype |  |  |  | 0.005 |
| ADHD, Other or Unspecified | 48 (9.9%) | 63 (18.6%) | 111 (13.5%) |  |
| ADHD, Combined subtype | 268 (55.4%) | 154 (45.4%) | 422 (51.3%) |  |
| ADHD, predominantly<br>hyperactive impulsive<br>subtype | 33 (6.8%) | 12 (3.5%) | 45 (5.5%) |  |
| ADHD, predominantly<br>inattentive subtype | 135 (27.9%) | 110 (32.4%) | 245 (29.8%) |  |
| Observational period (days) |  |  |  | <0.001 |
| Median (Q1, Q3) | 366<br>(196, 366) | 219<br>(99.5, 366) | 340<br>(153, 366) |  |
| Number of ADHD visits |  |  |  | <0.001 |
| Median (Q1, Q3) | 13.0 (7.0, 24.0) | 4.0 (4.0, 7.3) | 8.0 (4.0, 19.0) |  |

### Time to medication prescription

The median time from first ADHD diagnosis to first ADHD medication prescription was 4 days (interquartile range: 0-65 days), with a mean of 51 days. **Figure 2** illustrates the cumulative incidence of medication prescription within 12 months of diagnosis, stratified by patient race/ethnicity. Overall, 25% (n=202) of patients were prescribed medications at the first ADHD diagnostic visit. Throughout the first year after diagnosis, White patients showed the highest ADHD prescription rates, followed by Hispanic, Black, and Asian patients. At 12 months post-diagnosis, 64.6% of White patients received a medication prescription, followed by 58.0% of Hispanic, 50.0% of Black and 46.2% of Asian patients.

**Figure 2.**
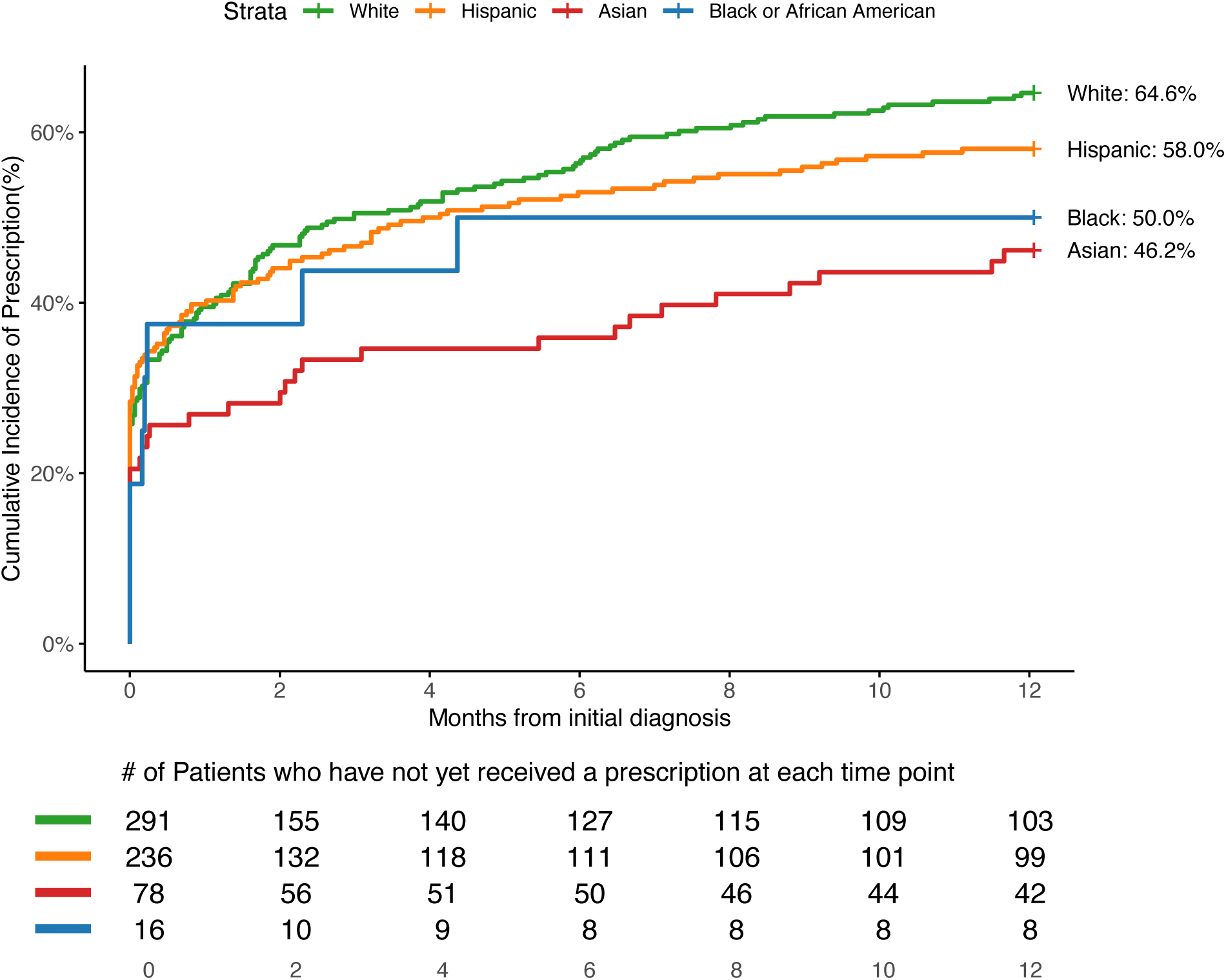
Cumulative Incidence of Prescription within 12 months, stratified by race/ethnicity.

### Regression model

**Table 2** shows the results of a Cox regression model that assessed the associations between patient predisposing, enabling, and need factors and time to ADHD medication prescription. Longer time to ADHD medication prescription was associated with Asian race/ethnicity (predisposing) (HR, 0.65, CI, 0.46-0.94), Psychologist clinician (enabling), (HR, 0.47, CI 0.32-0.68), as well as 1-2 comorbidities (HR, 0.76, CI, 0.61-0.94) and ≥3 comorbidities (HR, 0.64, CI, 0.47-0.89) (need). Shorter time to prescription was associated with older patient age (HR, 1.2, CI, 1.1-1.3) (predisposing) as well as the ADHD combined subtype (HR, 1.2, CI, 1.1-1.3) and ADHD hyperactivity/impulsivity subtype (HR 1.7, CI, 1.2-2.6) (need). Patient sex (predisposing) and the insurance type (enabling) were not statistically significantly associated with time to prescription of ADHD medication.

**Table 2.** Cox regression model (N=823): Associations between patient predisposing, enabling, and need factors and time to ADHD medication prescription.

| Predictors | Hazards Ratio | 95% Confidence Interval |  | P value |
| --- | --- | --- | --- | --- |
|  |  | Lower limit | Upper limit |  |
| **Predisposing** |  |  |  |  |
| Male vs Female | 1.017 | 0.821 | 1.260 | 0.878 |
| Age at diagnosis | 1.197 | 1.122 | 1.277 | <0.001 |
| Asian vs White | 0.653 | 0.456 | 0.936 | 0.020 |
| Black or African American vs White | 0.803 | 0.394 | 1.640 | 0.548 |
| Hispanic vs White | 0.912 | 0.716 | 1.162 | 0.456 |
| Other vs White | 0.412 | 0.131 | 1.299 | 0.130 |
| Unknown vs White | 0.904 | 0.709 | 1.153 | 0.415 |
| **Enabling** |  |  |  |  |
| Public vs Private Insurance | 0.989 | 0.796 | 1.230 | 0.921 |
| Nurse Practitioner vs Physician | 0.9144 | 0.5336 | 1.567 | 0.745 |
| Psychologist vs Physician | 0.4655 | 0.318 | 0.681 | <0.001 |
| **Need** |  |  |  |  |
| 1-2 Comorbidities vs none | 0.758 | 0.613 | 0.938 | 0.011 |
| ≥3 Comorbidities vs none | 0.644 | 0.466 | 0.900 | 0.008 |
| ADHD unspecified or others vs ADHD predominantly inattentive subtype | 0.753 | 0.539 | 1.051 | 0.096 |
| ADHD combined subtype vs ADHD predominantly inattentive subtype | 1.377 | 1.111 | 1.706 | 0.003 |
| ADHD predominantly hyperactive impulsive subtype vs ADHD predominantly inattentive subtype | 1.7314 | 1.1661 | 2.571 | 0.007 |

## DISCUSSION

In this retrospective cohort study, 59% of school-age children seen in DBP subspecialty clinics received ADHD medication prescriptions within one year of their first ADHD diagnostic visit. Among patients who received medication prescriptions, Asian race/ethnicity was associated with longer time to prescription. ADHD diagnostic visits conducted by a psychologist also were associated with a longer time to prescription. In contrast, and different from our initial hypothesis, having no comorbidities was associated with shorter time to prescription. Older patient age and certain ADHD subtypes (i.e., combined and predominantly hyperactive-impulsive presentations) were associated with shorter time to prescription, which fit our hypothesis.

When we compared the subgroup of patients who were prescribed an ADHD medication within a year of diagnosis to those who were not prescribed medications, we found that prescribed patients had more frequently an ADHD combined subtype diagnosis and were less frequently diagnosed by a psychologist. Patient sex, race/ethnicity, and insurance type did not differ across patients with and without medication prescription. These findings largely align with previous studies that examined patient and clinical factors associated with ADHD medication initiation. One population-based study of young adults in Sweden found older age and male sex were positively associated with medication initiation, whereas having certain comorbid conditions (e.g., depression and anxiety) was negatively associated with medication initiation.^17^ A study in a pediatric primary care setting also found that male sex was associated with medication receipt whereas patient race, comorbid conditions, and physician characteristics made no difference.^18^ Our study population represents patients seeking care in DBP subspecialty clinics, which differs from the general population and could explain why some of our findings are different from prior studies. Further studies in DBP settings are needed to assess the generalizability of our study findings.

When examining predisposing sociodemographic patient factors associated with time from diagnosis to prescription, we hypothesized that non-White children will have a longer time to initial prescriptions given prior literature that reported treatment delays in these sub-populations.^19^ Our findings supported this hypothesis and further highlighted that Asian race/ethnicity was most significantly associated with lower rates of and longer time to medication prescription compared to White patients. This finding is an important contribution to the literature because most studies that examined sociodemographic disparities in ADHD care focused on African American and Latinx populations.^20,21^ To further examine the low rates of medication prescription among Asian patients in our practice, we recently implemented a more granular documentation of race in patients referred to our clinical practice, disaggregating the Asian population into subgroups (e.g., Asian Indian, Chinese, Japanese). As a next step, we plan to combine quantitative and qualitative methods to uncover the drivers of variation in ADHD care in this under-studied population.

We also examined the possible contribution of patient age and sex, which are other predisposing factors. We found that older age, but not patient sex, was associated with shorter time to medication prescription. Other studies found that male patients were less likely to start ADHD medications.^13,15,18^ However, these studies did not examine the association between these factors and the <u>timing</u> of medication initiation after diagnosis.

When examining enabling factors associated with time from diagnosis to prescription, we found that an initial ADHD diagnosis by a psychologist was associated with longer time to medication prescription. We are not aware of prior studies that examined the influence of provider type in DBP settings on timing of medication treatment. Developmental-Behavioral Pediatrics clinics, in which school-age children are frequently diagnosed with ADHD, are unique in that along with physicians and nurse practitioners, psychologists can evaluate for and give the medical diagnosis of ADHD. The longer time to prescription likely relates to the fact that psychologists cannot prescribe medications. This is a confounding factor since an initial diagnosis by a psychologist means a patient must wait for a medical visit with a physician or nurse practitioner irrespective of desire to promptly start ADHD medication. This finding should be considered when planning which patients in a DBP practice will be first seen by a psychologist versus a prescribing clinician.

When examining need factors associated with time from diagnosis to prescription, we hypothesized that more comorbidities would result in shorter time to prescriptions. However, different from our initial hypothesis, having no comorbidities was associated with shorter time to prescription. A retrospective EHR study of primary care clinics found no association between comorbidities and rate of medication prescriptions in children with ADHD.^18^ On the other hand, a retrospective EHR study of preschool-age children with ADHD seen in DBP clinics showed that having multiple comorbid conditions was associated with an increased chance of being prescribed non-stimulant medications.^23^ It is possible that DBP clinicians in the present study chose to first address patient comorbidities before starting ADHD medication treatment, as suggested by one study of adult ADHD patients.^24^ Future studies are needed to evaluate factors influencing DBP clinician decision-making when prescribing ADHD medications to school-age children with ADHD who have multiple comorbidities.

Overall, use of the Andersen Health Care Utilization model allowed us to highlight associations between predisposing, enabling, and need factors and the timing of ADHD medication prescription. These results show that certain unique factors affect time to treatment of children with ADHD. Targeting these predictors by addressing the barriers at the time of the initial diagnostic visit could enhance timely treatment of ADHD and lead to better overall outcomes.

### Strengths and Limitations

This study has several strengths including the use of real-world clinical data from a large DBP practice, the large cohort size, and the diverse racial/ethnic backgrounds of cohort patients, including 29% Hispanic and 10% Asian patients. The use of a well-researched and well-established conceptual framework to capture health-care utilization in terms of ADHD is another important strength of the study.

There are also several study limitations. This study analyzed the records of a single subspecialty practice, which decreases the generalizability of the results. Another limitation is that this study only considered structured EHR data, without examination of the free text of patient notes. This limited our ability to examine more nuanced information about why families or clinicians did not start ADHD medications or what is affecting the time to start medication. Further studies are needed to understand both clinical and family factors that influence medication initiation. Lastly, this study did not address two major global and national events that occurred during the timespan of this study. We did not specifically address the COVID-19 pandemic nor the national shortage of stimulant medications, which may have affected time from diagnosis to prescription.^25,26^

## CONCLUSION

This retrospective cohort study contributes to the limited literature on factors that influence the timing of ADHD medication treatment in DBP practice. The results of this study reinforce the fact that both sociodemographic and clinical factors influenced the initiation of ADHD medication in school-age children seen in the examined DBP practice. Our results further suggest that patient race/ethnicity, specifically Asian, affect time to medication treatment. This warrants further studies to understand how family cultural factors may explain these findings. In addition, the presence of multiple comorbidities was a deterrent to early medication treatment. Future mixed-methods studies are needed to evaluate family, clinician, and system factors that influence the shared decision of families and DBP clinicians on the timing of medication initiation in school-age children with ADHD.

## Supporting information

eSupplement 1

## Data Availability

The dataset generated and analyzed in the current study contains protected patient health information and is therefore not publicly available; the data will be shared on reasonable request to the corresponding author.

## Acknowledgments

This research used data or services provided by STARR, “STAnford medicine Research data Repository,” a clinical data warehouse containing live Epic data from Stanford Health Care (SHC), the Stanford Children’s Hospital (SCH), the University Healthcare Alliance (UHA) and Packard Children’s Health Alliance (PCHA) clinics and other auxiliary data from Hospital applications such as radiology PACS. STARR platform is developed and operated by Stanford Medicine Research Information Technology (IT) team and is made possible by Stanford School of Medicine Research Office. We thank Stanford Medicine Research IT for their support and assistance in data acquisition and extraction.

## REFERENCES

1. Li Y, Yan X, Li Q, et al. Prevalence and Trends in Diagnosed ADHD Among US Children and Adolescents, 2017-2022. JAMA Netw Open. 2023;6(10):e2336872. doi:10.1001/jamanetworkopen.2023.36872

2. Sibley MH, Arnold LE, Swanson JM, et. al. Variable Patterns of Remission from ADHD in the Multimodal Treatment Study of ADHD. AJP. 2022;179(2):142–151. doi:10.1176/appi.ajp.2021.21010032

3. Yoshimasu K, Barbaresi WJ, Colligan RC, et al. Adults With Persistent ADHD: Gender and Psychiatric Comorbidities-A Population-Based Longitudinal Study. J Atten Disord. 2018;22(6):535–546. doi:10.1177/1087054716676342

4. Charach A, Yeung E, Climans T, et al. Childhood Attention-deficit/hyperactivity disorder and Future Substance Use Disorders: comparative meta-analyses. J Am Acad Child Adolesc Psychiatry. 2011;50(1):9–21. doi:10.1016/j.jaac.2010.09.019

5. Curry AE, Metzger KB, Pfeiffer MR, et al. Motor Vehicle Crash Risk Among Adolescents and Young Adults With Attention-Deficit/Hyperactivity Disorder. JAMA Pediatr. 2017;171(8):756–763. doi:10.1001/jamapediatrics.2017.0910

6. Storebø OJ, Storm MRO, Pereira Ribeiro J, et al. Methylphenidate for Children and Adolescents with Attention Deficit Hyperactivity Disorder (ADHD). Cochrane Database Syst Rev. 2023;3(3):CD009885. doi:10.1002/14651858.CD009885.pub3

7. Biederman J, Monuteaux MC, Spencer T, et al. Do stimulants protect against psychiatric disorders in youth with ADHD? A 10-year follow-up study. Pediatrics. 2009;124(1):71–78. doi:10.1542/peds.2008-3347

8. Harpin V, Mazzone L, Raynaud JP, et al. Long-Term Outcomes of ADHD: A Systematic Review of Self-Esteem and Social Function. J Atten Disord. 2016;20(4):295–305. doi:10.1177/1087054713486516

9. Wolraich ML, Hagan JF, Allan C, et al. Clinical Practice Guideline for the Diagnosis, Evaluation, and Treatment of Attention-Deficit/Hyperactivity Disorder in Children and Adolescents. 2019;144(4).

10. Barbaresi WJ, Campbell L, Diekroger EA, et al. Society for Developmental and Behavioral Pediatrics Clinical Practice Guideline for the Assessment and Treatment of Children and Adolescents with Complex Attention-Deficit/Hyperactivity Disorder. J Dev Behav Pediatr. 2020;41(2S):S35–S57. doi:10.1097/DBP.0000000000000770

11. Dodds M, Wanni Arachchige Dona S, Gold L, et al. Economic Burden and Service Utilization of Children With Attention-Deficit/Hyperactivity Disorder: A Systematic Review and Meta-Analysis. Value in Health. 2024;27(2):247–264. doi:10.1016/j.jval.2023.11.002

12. Danielson ML, Bitsko RH, Ghandour RM, et al. Prevalence of Parent-Reported ADHD Diagnosis and Associated Treatment Among U.S. Children and Adolescents, 2016. Journal of Clinical Child & Adolescent Psychology. 2018;47(2):199–212. doi:10.1080/15374416.2017.1417860

13. Morgan PL, Hu EH. Sociodemographic Disparities in ADHD Diagnosis and Treatment among U.S. Elementary Schoolchildren. Psychiatry Research. 2023;327:115393. doi:10.1016/j.psychres.2023.115393

14. McKenna K, Wanni Arachchige Dona S, Gold L, et al. Barriers and Enablers of Service Access and Utilization for Children and Adolescents With Attention Deficit Hyperactivity Disorder: A Systematic Review. J Atten Disord. 2024;28(3):259–278. doi:10.1177/10870547231214002

15. Pastor PN, Simon AE, Reuben CA. ADHD: Insurance and Mental Health Service Use. Clin Pediatr (Phila). 2017;56(8):729–736. doi:10.1177/0009922816673401

16. Lederle M, Tempes J, Bitzer EM. Application of Andersen’s behavioural model of health services use: a scoping review with a focus on qualitative health services research. BMJ Open. 2021;11(5):e045018. doi:10.1136/bmjopen-2020-045018

17. Gémes K, Taipale H, Björkenstam E, et al. The Role of Sociodemographic and Clinical Factors in the Initiation and Discontinuation of Attention Deficit Hyperactivity Disorder Medication among Young Adults in Sweden. Front Psychiatry. 2023;14:1152286. doi:10.3389/fpsyt.2023.1152286

18. Kamimura-Nishimura KI, Epstein JN, Froehlich TE, et al. Factors Associated with Attention Deficit Hyperactivity Disorder Medication Use in Community Care Settings. The Journal of Pediatrics. 2019;213:155–162.e1. doi:10.1016/j.jpeds.2019.06.025

19. Shi Y, Hunter Guevara LR, Dykhoff HJ, et al. Racial Disparities in Diagnosis of Attention-Deficit/Hyperactivity Disorder in a US National Birth Cohort. JAMA Netw Open. 2021;4(3):e210321. doi:10.1001/jamanetworkopen.2021.0321

20. Kamimura-Nishimura K, Bush H, Amaya De Lopez P, et al. Understanding Barriers and Facilitators of Attention-Deficit/Hyperactivity Disorder Treatment Initiation and Adherence in Black and Latinx Children. Academic Pediatrics. 2023;23(6):1175–1186. doi:10.1016/j.acap.2023.03.014

21. Jhawar N, Antshel K. Asian Indian American Parental Help-Seeking Intentions for ADHD. Res Child Adolesc Psychopathol. 2023;51(11):1551–1563. doi:10.1007/s10802-023-01108-2

22. Blum NJ, Shults J, Harstad E, et al. Common Use of Stimulants and Alpha-2 Agonists to Treat Preschool Attention-Deficit Hyperactivity Disorder: A DBPNet Study. J Dev Behav Pediatr. 2018;39(7):531–537. doi:10.1097/DBP.0000000000000585

23. Katzman MA, Bilkey TS, Chokka PR, et al. Adult ADHD and Comorbid Disorders: Clinical Implications of a Dimensional Approach. BMC Psychiatry. 2017;17(1):302. doi:10.1186/s12888-017-1463-3

24. Bannett Y, Dahlen A, Huffman LC, et al. Primary Care Diagnosis and Treatment of Attention-Deficit/Hyperactivity Disorder in School-Age Children: Trends and Disparities During the COVID-19 Pandemic. J Dev Behav Pediatr. 2022;43(7):386–392. doi:10.1097/DBP.0000000000001087

25. He S, Esteban McCabe S, Conti RM, et al. Prescription Stimulant Dispensing to US Children: 2017–2023. Pediatrics. 2025;155(2):e2024068558. doi:10.1542/peds.2024-068558

