## Supplementary material for "Time to Medication in School-Age Children with ADHD: Assessing the Effect of Sociodemographic and Clinical Factors": eSupplement 1

**eSupplement.** List of codes used for comorbid conditions.

| **Condition type** | **ICD-10 code** | **Concept id*** | **Condition name** |
| --- | --- | --- | --- |
| Autism | F84.0 | 35207254 | Autism spectrum disorder |
| Autism | F84.5 | 35207257 | Asperger's syndrome |
| Autism | F84.8 | 35207258 | Other pervasive developmental disorders |
| Autism | F84.9 | 35207259 | Pervasive developmental disorder, unspecified |
| Autism | F94.8 | 35207278 | Other childhood disorders of social functioning |
| Autism | F94.9 | 35207279 | Childhood disorder of social functioning, unspecified |
| Autism | F80.82 | 37200331 | Social pragmatic communication disorder |
| Anxiety | F40 | 45595906 | Phobic anxiety disorders |
| Anxiety | F41 | 45571763 | Other anxiety disorders |
| Anxiety | F93.0 | 35207272 | Separation anxiety disorder of childhood |
| Anxiety | F93.8 | 35207273 | Other childhood emotional disorders |
| Anxiety | F93.9 | 35207274 | Childhood emotional disorder, unspecified |
| Anxiety | F94.0 | 35207275 | Selective mutism |
| Anxiety | R45.0 | 35211360 | Nervousness |
| Anxiety | R45.82 | 45568135 | Worries |
| Depression | F32 | 45595904 | Depressive episode |
| Depression | F33 | 45576542 | Recurrent depressive disorder |
| Depression | F34 | 45557203 | Persistent mood [affective] disorder |
| Depression | F39 | 45591137 | Unspecified mood [affective] disorder |
| Depression | R45.86 | 45539347 | Emotional lability |
| Depression | R45.81 | 45539346 | Low self-esteem |
| Sleep problems | G47 | 45576588 | Sleep disorders |
| Sleep problems | F51 | 45552491 | Sleep disorders not due to a substance or known physiological condition |
| Sleep problems | Z72.82 | 1576169 | Problems related to sleep |
| Sleep problems | Z73.81 | 45537670 | Behavioral insomnia of childhood |
| Behavioral problem | F91 | 45600764 | Conduct disorders (including oppositional defiant disorder) |
| Behavioral problem | F63 | 45576555 | Habit and impulse disorders |
| Behavioral problem | Z72.810 | 45581064 | Child and adolescent antisocial behavior, behavior problem at school |
| Behavioral problem | R45.4 | 35211364 | Irritability and anger |
| Behavioral problem | R45.5 | 35211365 | Hostility |
| Behavioral problem | R45.6 | 35211366 | Violent behavior |
| Learning problem/ disability | F81 | 45591156 | Specific developmental disorders of scholastic skills |
| Learning problem/ disability | Z55 | 45552089 | Problems related to education and literacy |
| Learning problem/ disability | R48.0 | 35211378 | Dyslexia and alexia |
| Language delay/ disorder | F80.0 | 35207244 | Phonological disorder |
| Language delay/ disorder | F80.1 | 35207245 | Expressive language disorder |
| Language delay/ disorder | F80.2 | 35207246 | Mixed receptive-expressive language disorder |
| Language delay/ disorder | F80.81 | 45591155 | Childhood onset fluency disorder |
| Language delay/ disorder | F80.89 | 45538086 | Other developmental disorders of speech and language |
| Language delay/ disorder | F80.9 | 35207249 | Developmental disorder of speech and language, unspecified |
| Language delay/ disorder | R47.89 | 45563311 | Other speech disturbances |
| Language delay/ disorder | R47.9 | 45539349 | Unspecified speech disturbances |
| Global Developmental Delay/Intellectual disability | F88 | 35207260 | Global Developmental Delay/Other disorders of psychological development |
| Global Developmental Delay/Intellectual disability | F89 | 35207261 | Unspecified disorder of psychological development |
| Global Developmental Delay/Intellectual disability | F70 | 35207238 | Mild intellectual disabilities |
| Global Developmental Delay/Intellectual disability | F71 | 35207239 | Moderate intellectual disabilities |
| Global Developmental Delay/Intellectual disability | F72 | 35207240 | Severe intellectual disabilities |
| Global Developmental Delay/Intellectual disability | F73 | 35207241 | Profound intellectual disabilities |
| Global Developmental Delay/Intellectual disability | F79 | 35207243 | Unspecified intellectual disabilities |

*We included these concept ids and all their descendants
